# Heterogeneity in referral preferences of women at high risk for postpartum depression: a discrete choice experiment

**DOI:** 10.64898/2026.03.25.26349110

**Authors:** Xin Jin, Lauren Zhang, Hui Li, Wenjie Gong

**Affiliations:** HER Team and Department of Maternal and Child Health, Xiangya School of Public Health, Central South University, Hunan, 410078, China; Department of Community Healthcare, Kunshan Maternity and Children’s Health Care Hospital, Jiangsu, 215300, China; Columbia University Vagelos College of Physicians and Surgeons, New York, New York, United States of America; School of Mathematics and Statistics, University of Birmingham, Birmingham, B15 2TT; Xiangya School of Public Health, Central South University, 238 shangmayuanling, Xiangya Rd, Kaifu District, Changsha, Hunan, 410078, China

**Author notes:** Xin Jin, MD, Wenjie Gong,.

## Abstract

Despite the global prevalence of postpartum depression (PPD), current referral uptake rates are far from satisfactory. While some qualitative studies have investigated factors affecting PPD referrals, a gap in quantitative analysis remains. Addressing this, our study utilized a discrete choice experiment (DCE) to understand the procedural elements influencing PPD referral uptake among diagnosed women. The DCE was conducted via home visits by healthcare providers and a comprehensive mobile app questionnaire. We constructed seven distinct referral attributes to explore participants’ preferences, analyzed using mixed logit models and latent class analysis. This analysis identified key determinants and revealed the heterogeneities in referral preferences. A total of 698 individuals completed the DCE questionnaire. All assessed attributes, except for Accompaniment (going to clinic with a family member), were important determinants of preference. Participants generally preferred referrals to psychiatric clinics, face-to-face consultations, lower costs, and shorter waiting times. Significantly, participants’ personal and socio-demographic characteristics also played a critical role in their referral preferences. Latent class analysis categorized participants into four distinct groups based on their preferences, with treatment cost and waiting times being the most decisive factors. In conclusion, the preference for PPD referrals is predominantly driven by convenience and access to specialist care. To enhance referral uptake, developing flexible and personalized referral programs that cater to these preferences is crucial.

## 1. Introduction

China is currently undergoing a significant transformation in its epidemiology and demographics, shifting its focus from primarily reducing maternal and child mortality rates to providing high-quality healthcare services (Qiao et al., 2021). In 2020, China launched an ambitious plan to enhance maternal mental health (Commission, 2020). This initiative includes routine screening for perinatal depression and advocates for strengthened professional support to improve maternal mental well-being. A systematic review reported that the positive rate of postnatal depression (PPD) screening in mainland China is 14.8% (95% C.I. 13.1%, 16.6%) and 16.3% (95% C.I. 14.7%,18.2%) for perinatal depression (Nisar et al., 2020). However, despite efforts and recommendations aimed at enhancing mental health services and providing effective perinatal care for women, the referral uptake rates for PPD remain low. For example, in a cohort study involving 1,126 women in Hunan, China, the referral rate was only 0.4% (Gong et al., 2020). Similarly, the uptake of referrals among PPD patients in other countries is also limited, with only 43% of women with positive screening results accepting referrals, even in developed economies, according to a meta-analysis (W Xue et al., 2020). This low referral uptake underscores the significance of PPD as a major public health challenge (Hahn-Holbrook et al., 2017), and emphasizes the need to establish more engaging referral programs that are more likely to be accepted by PPD patients. Ultimately, the effectiveness of routine PPD screening largely depends on the willingness of women who test positive for PPD to engage in follow-up referrals..

At present, most existing studies used qualitative research methods to explore the factors that affect the referral of PPD qualitatively (Elkhodr et al., 2018; Flynn et al., 2010; Fonseca et al., 2015; Holopainen, 2002; J. J. Kim et al., 2010). Drawing a broad reference, a meta-study by Forde et al analysed factors or barriers affecting recovery from postpartum psychosis and called for “an integrated and individualized” approach to improving clinical outcomes(Forde et al., 2020). In a most recent qualitative study on PND referrals in China, researchers have identified five areas such as individual, interpersonal, institutional, community and public policy that could affect new mothers accepting referrals(Xue et al., 2023). Building upon these studies, it is therefore necessary to evaluate the factors that affect the referral of PPD to establish a set of effective referral programs.

Despite the wide spectrum of studies on PPD, there is a dearth of research assessing patients’ preferences for accepting referrals. To address this gap, the objective of this study is to elicit and analyze the stated preferences of Chinese PPD patients for referrals quantitatively using the discrete choice experiment (DCE), which is regarded as the preferred method(Aristides et al., 2004; Barber et al., 2019; Fawsitt et al., 2017; Fegert et al., 2011; Karns & Khera, 2015; King et al., 2007; Minton et al., 2017; Sculpher et al., 2004). DCE assumes that patients can reveal their preferences by making choices based on the characteristics of attributes(de Bekker-Grob et al., 2019).^25^ Aligning treatment with patients’ preferences could lead to better treatment adherence and enhanced efficiency in mental health care. Tünneßen et al reviewed eleven existing DCEs concerning anxiety and depressive disorders and found that patients considered the attributes of the referral process itself and the cost to be more significant than the treatment outcome; clinicians and policymakers should take note of these attributes and levels(Tünneßen et al., 2020). Building on the literature, we created a series of choice sets for referral uptake with varying levels of attributes, along with an opt-out option, and demographic information for each patient. As a result, each PPD patient would be presented with various combinations of referral options, enabling us to examine patients’ real-world preferences for each attribute. By developing a mechanism that meets the varied preferences of PPD patients, healthcare managers have the potential to significantly improve referral uptake rates.

## 2. Methods

### 2.1 Study sample and data collection

In the study, we employed a stratified cluster sampling approach to obtain a sample representative of the population in Changsha, China. Specifically, there are seven districts and counties managed by Changsha municipal city. Out of the seven districts and counties, we randomly selected two which resulted in Changsha County and Kaifu District. All the maternal healthcare providers in the two districts and all the pregnant women under their care who needed a postpartum visit were included in our sample collection.

Maternal healthcare providers are medical professionals specializing in maternal health management within primary health care in China. They frequently interact with pregnant women during the perinatal period. As part of the national Public Health Service, these providers monitor both the mother’s and newborn’s health parameters, such as blood pressure and weight. All the healthcare providers in our study had received specialized training to administer the questionnaire, which included explaining the importance of the maternity program and the process of obtaining informed consent.

Participants completed the questionnaire using their mobile phones during the home visits conducted by the maternal healthcare providers. Initially, participants filled out a questionnaire assessing their risk of PPD. Instantly upon completion, EPDS scores were calculated. Those who scored ten or above on the EPDS, indicating a positive screen for PPD, were automatically directed to the subsequent DCE questionnaire. Consent forms for the study and survey were signed by all participants at the onset of the survey. The full survey process is illustrated in Figure 1. The study received approval from the Institutional Review Board at Central South University (CTXY-2020-102).

**Figure 1.**
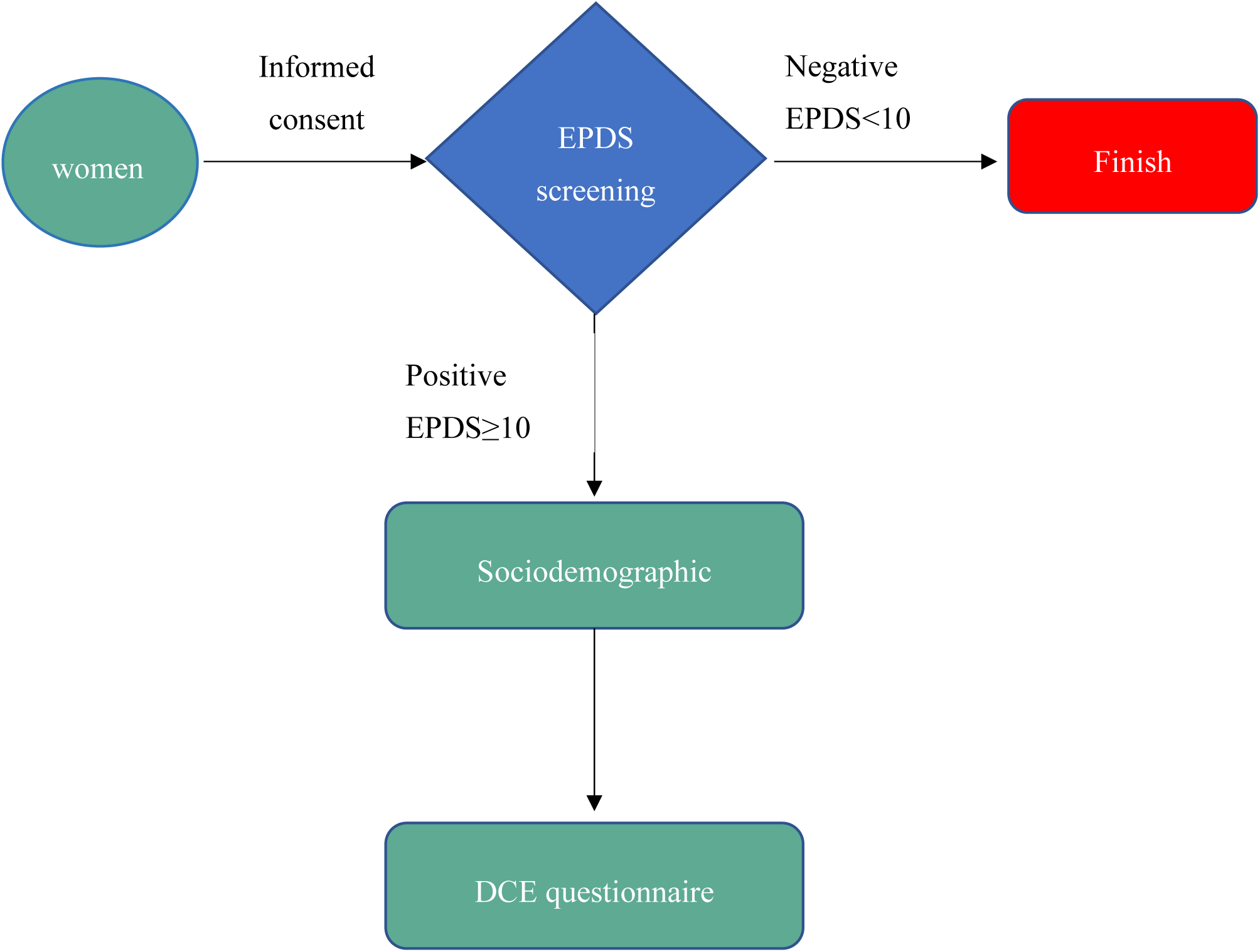
Flow chart of survey

Of the 2,134 new mothers who were discharged from the hospital within seven days post-delivery, 617 were found to have screened positive for PPD (with an EPDS score of ten or above) based on home visit interviews. The minimum required sample size was determined using the methods of de Bekker-Grob (de Bekker-Grob et al., 2015). To account for an estimated opt-out rate of 25%, we aimed to recruit at least 522 participants to ensure an adequately sized sample.

### 2.2 Discrete Choice Experiment

In our study utilizing a DCE approach, we hypothesize that the choices PPD patients make about referrals are influenced by various characteristics or attributes. These include the referral route, referral initiator, referral destination, interval time, being accompanied, consultation length and expenses. Participants are presented with a range of options and prompted to indicate their preferred PPD referral choice from these predefined sets of levels. For example, in terms of referral interval times, participants could select intervals such as one hour, six hours, twelve hours, or twenty-four hours. We subsequently analyze the trade-offs they make when selecting from the diverse combinations of levels and attributes concerning PPD referrals.

#### 2.2.1 Attributes and Attribute Levels

Attributes and their respective levels are essential components in DCE studies. In our study, these attributes were determined through various sources. Firstly, we conducted a qualitative study based on participant observations and interviews in primary care settings in China, building upon a previous qualitative study by Xue et al. (2023). Four investigators conducted 30-day participant observations in four primary health centers, both in urban and rural areas. Data were collected through participant observations and semi-structured, in-depth interviews with new mothers who had positive results on postnatal depression screening, their family members, and primary healthcare providers. From this study, we selected key attributes and potential levels initially.

Subsequently, we identified the attributes and their corresponding levels through a literature review of articles related to PPD referrals and conducted focus group interviews. This was followed by discussions with eight experts, including two psychiatrists, one senior maternal health researcher, and four qualitative interview administrators. In the focus group study, we conducted individual interviews with seventeen pregnant women who had scored more than ten points on the EPDS; among them, two had previously been diagnosed with PPD and had undergone standard treatment in psychiatric clinics. Interview data analysis was guided by grounded theory principles (Elliott & Jordan, 2010; Glaser & Strauss, 2008; Strauss & Corbin, 1998). Initially, two-thirds of the text were analyzed, after which another researcher independently analyzed the remaining content using the same methodology. The outcomes of both analyses were compared to achieve theoretical saturation(Francis et al., 2010).

Following this analysis, the experts were invited back to discuss and modify the original attributes. Closely following the insights from the qualitative interviews and clinical practice, we then defined the levels. For example, the structure expense in the questionnaire closely followed the actual consultation fees charged by tertiary hospitals in Changsha. The levels of referral waiting time were also adopted from the waiting hours for patients. Finally, we carried out a polite study to validate the levels in the attributes. The final attributes and levels included in the DCE are presented in Table 1.

**Table 1.** The attributes and levels of referral preference.

| Attribute | Level | Explanation |
| --- | --- | --- |
| Referral initiator | Obstetrician (base level)<br>Maternal healthcare officer | The person initiating the referral. This term describes whether the role of medical professionals makes a difference. This attribute was derived from Xue et al. (2023), the expert panel, and focus group discussions. During pregnancy and the delivery period, women in China will regularly meet with their maternal healthcare officers and will consult their obstetrician if needed. |
| Referral destination | Maternal clinic (base level)<br>Psychiatric clinic | The department receiving referrals. For mental healthcare in China, women can choose either to visit a maternal clinic or psychiatric clinic if they get a referral. This attribute originated from Xue et al. (2023), the expert panel and focus group discussions. |
| Referral route | Online (base level)<br>In-person | Whether referral is carried out in person or via internet call (online). Given that the data collection for this study occurred during the COVID-19 pandemic, both in-person and online referrals were considered. Online consultations were particularly in demand during this period. The inclusion of this attribute was based on Xue et al. (2023), the expert panel, and discussions within the focus group. |
| Referral waiting time | One hour<br>Six hours<br>Twelve hours<br>Twenty-four hours (base level) | The time taken from accepting the referral to starting the consultation appointment, including travel time. We drew information from Xue et al. (2023) and the focus group. We observed and collected information in clinics on referral waiting time, which was usually up to 24 hours during the data collection period. The levels were validated in our pilot study. |
| Accompaniment | With family<br>Alone (base level) | Whether the mental care facility allows family members to be present. The inclusion of this attribute was based on Xue et al. (2023), the expert panel and discussions within the focus group. The two levels were validated in our pilot study. |
| Consultation length | One hour<br>Two hours (base level) | The time spent in the consultation clinic after the referral. We drew information from Xue et al. (2023), the expert panel and the focus group discussions. The consultation time and fees are usually set at a fixed hourly rate for convenience. The levels were validated in our pilot study. |
| Expenses (¥) | 100<br>300<br>500<br>700 | Full cost incurred over the course of referral and consultation. It has four levels, measured in hundreds, for convenience of comparison. This attribute originates from Xue et al. (2023); the levels were discussed by the expert panel and the focus group. We observed and collected information in clinics and hospitals; the consultation fees are structured at ¥100, ¥300, ¥500, and ¥700 and more, corresponding to different seniorities of consultants. The levels were validated in our pilot study. |

#### 2.2.2 Choice sets and questionnaire

Given the combination of seven attributes, each with two to four levels, there could be as many as 512 referral preference alternatives for a single participant. To mitigate an overwhelming cognitive burden on participants, this study employed orthogonal designs with foldover, constructing 16 pairs of referral choice sets (using a random seed of 221768). Each set presented two referral alternatives, displayed side-by-side for a paired comparison, with an opt-out alternative also provided, as shown in Figure 2. Our DCE development adhered to the International Society for Pharmacoeconomics Outcomes Research’s guidelines for conjoint analysis in health (Bridges et al., 2011). The DCE framework was constructed using SPSS 25.0.

**Figure 2.**
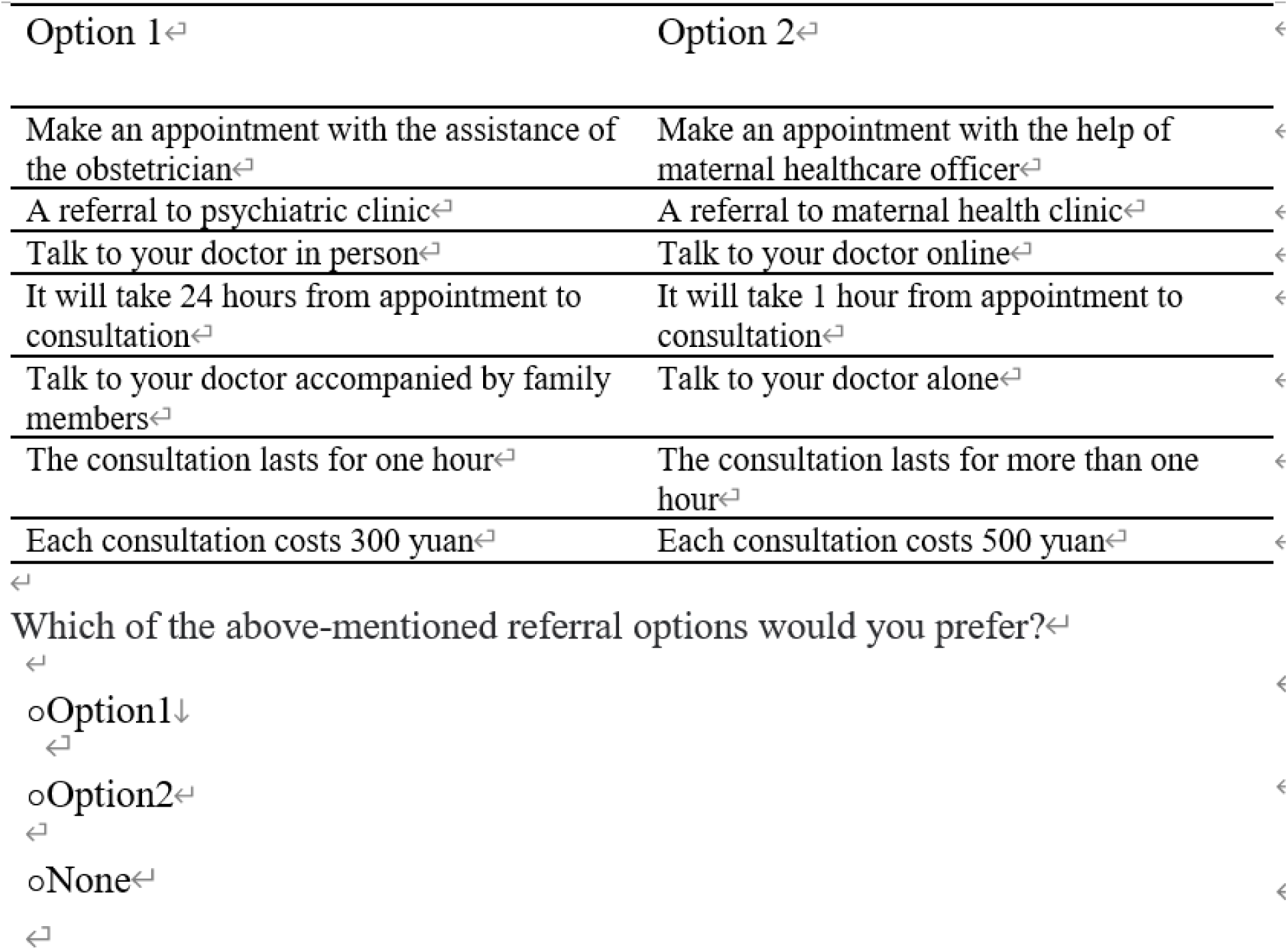
Example of a choice set

Every questionnaire commenced with an overview of the study’s objectives. A preliminary exercise was incorporated to acquaint respondents with the attributes, levels, and alternatives they would encounter. We implemented consistency checks using position change treatment to ensure genuine engagement and honest responses. The primary section of the DCE consisted of 16 choice sets, with each set offering two referral alternatives. Participants chose their preferred referral option in a manner similar to the example in Figure 2. Additionally, we recorded each participant’s Edinburgh Postnatal Depression Scale (EPDS) score (Costafreda et al., 2009; Meltzer-Brody et al., 2013), utilizing its Chinese translation for this study(Wang et al., 2009).After initially acquiring participants’ consent, we also gathered sociodemographic details such as age, residence, and occupation in the questionnaire. A pilot study, which involved 30 participants, was conducted to evaluate the questionnaire’s clarity and length.

The final survey questionnaire was programmed into So Jump (WenJuanXing), a prominent online survey platform in China. Participant scanned a QR code via the mobile app WeChat (Wei Xin) to access the questions within a week post-hospital discharge from the hospital. Spanning twelve screen-pages, the survey was designed for user-friendliness, allowing participants to easily navigate and hover over the attributes and levels on their mobile devices.

### 2.3 Statistical model

Our analysis is mainly based on the random utility model and latent class analysis. In this framework, an individual, denoted as n, chooses from j alternative choices. The choice is driven by the desire to maximize utility, which can be thought of as benefit or satisfaction. Specifically, individual n will opt for choice i over any other choice j if and only if *U*_*ni*_ > *U*_*nj*_, ∀*i* ≠ *j* ∈ *J*, where U is the utility for a given choice. The utility function (U) for individual n corresponding to choice i consisted of the observed utility level of m choice attributes, x1,…, xm, and a random error term *ɛ*_*ni*_. These attributes (x1,…, xm) included factors listed in Table 1 such as referral initiator, destination, route, interval length, accompaniment, consultation length and expenses.

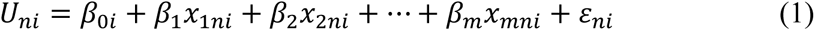

where *βs* are parameters associated with each attribute.

The mixed logit model was employed, allowing parameters to vary across individuals. These models were fitted using Stata 15.0 to obtain estimated coefficients and to infer the impact of attributes on individual choices (Lancsar & Louviere, 2008; Mangham et al., 2009). The mixed logit model also includes an alternative-specific constant (ASC) for referral alternatives. Participants’ characteristics, including sociodemographic factors, were analyzed as interactions with the ASC or other attributes. Mean willingness-to-pay (WTP) values were calculated by taking the expectations of the ratio of the coefficients of each referral attribute to the coefficient of cost. Their confidence intervals were subsequently estimated using the Delta method (Hole-Arne, 2013).

Under the assumption that preference heterogeneity has a discrete distribution, we further implemented latent class analysis with Mplus7.0. By allowing different parameters, this approach considers heterogeneity as indicative of the presence of various “potential” groups. When an individual n opts for the k^th^ option from all choices in scenario m, the utility of choosing category c is the largest. The utility function is as follows:

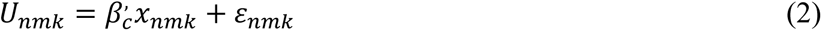

where *U*_*nmk*_ is the probability of individual n choosing alternative k in scenario m, *x*_*nmk*_ is the set of all attributes and characteristics displayed in the utility function, *β’_c_* stands for the group parameters and *ɛ*_*nmk*_ is the unobservable error.

In the latent class analysis, after examining the signs and significance of the fitted coefficients, we also calculated and evaluated the relative importance statistics for each attribute (Determann et al., 2016; Maaya et al., 2018; Ride & Lancsar, 2016; Vermunt & Magidson, 2005). This allowed us to observe the varying patterns across each class.

## 3. Results

### 3.1 Sample information

From December 1, 2020 to March 10, 2021, we collected a total of 2,134 screening questionnaires. Of these, 765 were from women suspected of having PPD with EPDS score greater than or equal to 10. Out of the 698 women who completed the subsequent referral preference survey, 81 did not pass the choice consistency test and were thus excluded from the analysis, resulting in a final sample of 617. That is, about 88% of participants exhibited stable preferences while answering the choice set question. Figure 3 exhibits a flowchart that led to the final sample. Using the AAPOR standardized calculation, the response rate was 92.1% (English, 2016). The socio-demographic information in the final sample was in general consistent with national, province or regional data, slightly fewer percentage women lived in the urban area 0.490 versus 0.572 in the regional data. Table 2 presented the summary statistics of participants characteristics.

**Figure 3.**
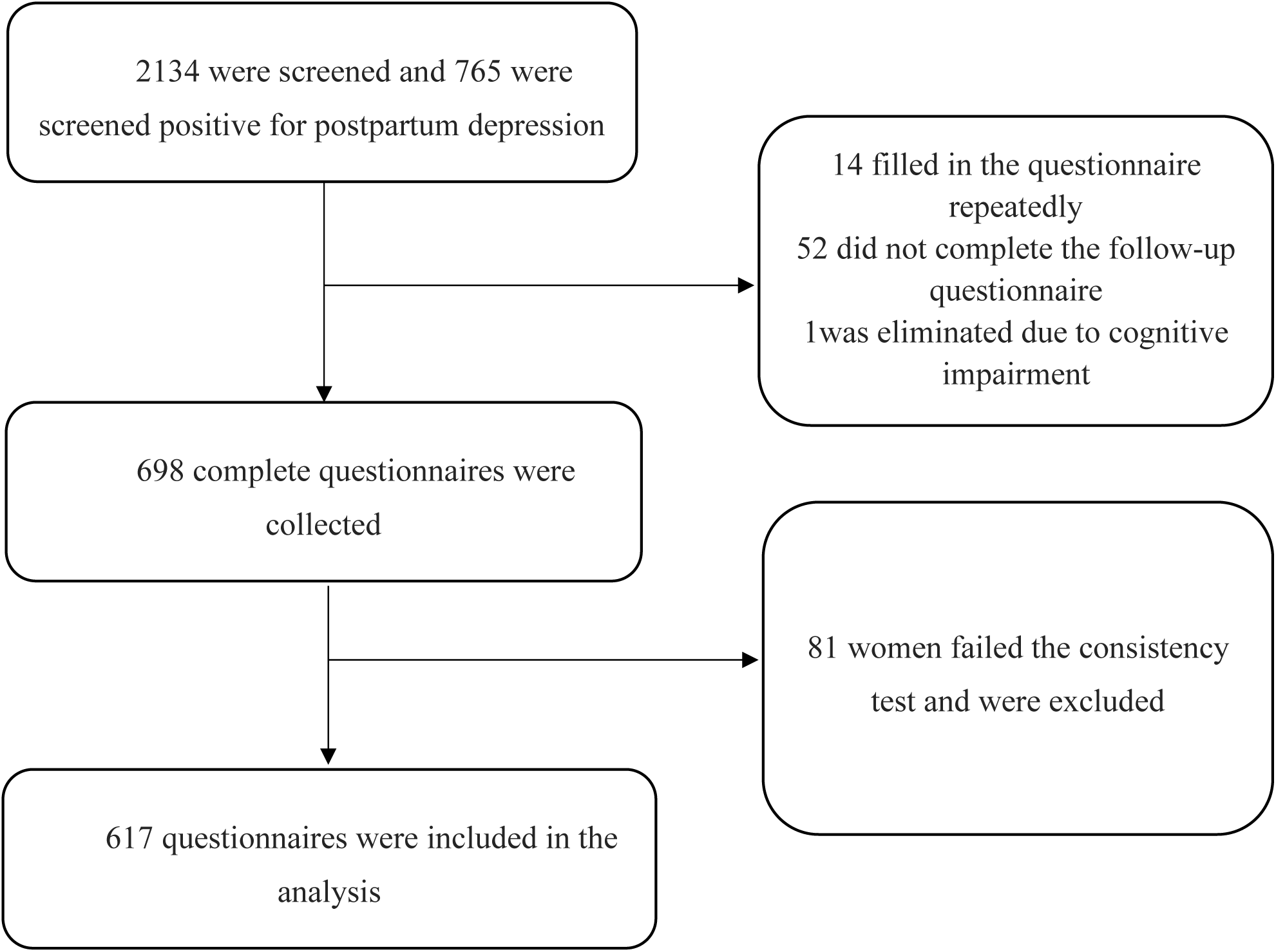
Flow chart of recruitment

**Table 2.** Descriptive statistics of participants characteristics [N=617].

| Variables | Description | Sample | National Data <sup>a</sup> |
| --- | --- | --- | --- |
|  |  | Mean<br>(SD) | [Province/Region]<br>Mean |
| EPDS | The score of Edinburgh Postnatal Depression Scale translated in Chinese | 12.796<br>(2.803) |  |
| Employed in public sector | Whether the participant was a public sector employee, yes=1, otherwise 0. | 0.118<br>(0.323) |  |
| Past depression | Whether the participant experienced depression before, yes=1, otherwise 0. | 0.047<br>(0.212) |  |
| Past miscarriage | Whether the participant experienced miscarriage before, yes=1, otherwise 0. | 0.267<br>(0.443) |  |
| Monthly income (CNY) | Monthly income of the participant in CNY (¥). | 7081<br>(3755) |  |
| Care by in-laws | Whether the participant was under care of her in-laws after delivery, yes=1, otherwise 0. | 0.214<br>(0.410) |  |
| Natural birth | Whether the participant just gave a natural birth to the new-born, yes=1, otherwise 0. | 0.571<br>(0.495) | 0.555<sup>b</sup><br>[0.542]<sup>b</sup> |
| Satisfied with the partner | Whether the participant was satisfied with her partner, yes=1, otherwise 0. | 0.856<br>(0.352) |  |
| Age | The age of the participant (in years). | 30.324<br>(4.387) | 29.1<sup>b</sup> |
| 19 < Age < 34 | Percentage of newborn mothers with age less than 34, greater than 19. | 0.902<br>(0.047) | 0.815<sup>b</sup><br>[0.831]<sup>b</sup> |
| College | Whether the participant had a bachelor or a postgraduate degree, yes=1, otherwise 0. | 0.412<br>(0.492) | 0.488<sup>b</sup><br>[0.444]<sup>b</sup> |
| Urban | Whether the participant lived in the urban area, yes=1, otherwise 0. | 0.402<br>(0.490) | 0.606<sup>c</sup><br>[0.572]<sup>c</sup> |

Notes: <sup>a</sup>: An empty cell means the data is not available for national, provincial, or regional populations.
<sup>b</sup>: Data comes from Annual Report on Maternal and Child Health Screening 2019.
<sup>c</sup>: Data comes from National Annual Statistics 2019.

On average, participants took about 20 minutes to complete the surveys. Their mean age was 30.3 years old (standard deviation=4.4). 72% of them resided in urban areas, 88.8% had at least a high school education, and 41.2% held an undergraduate degree or higher. The median monthly income was ¥5,000, and 55.6% of participants reported a decrease in monthly income after pregnancy. A significant majority (95.3%) had no recorded history of mental illness, 26.7% had suffered miscarriage in the past, and 20.1% experienced postpartum complications from their most recent pregnancy.

An EPDS score greater than 13 suggests that the mother is at a high risk of depression.^32^ We further divided participants into two groups: a non-high-risk group (EPDS score below thirteen) comprising 338 participants, and a high-risk group (EPDS score of thirteen or higher) with 279 participants. T-test comparisons revealed no significant differences between the groups regarding medical history or demographic and socioeconomic factors. The t-test results are provided in Appendix A.

### 3.2 DCE modelling results

Table 3 displays the results from the mixed logit models of referral preferences. The number of iterations was set at 500, with convergence achieved after 12 iterations. The attribute model presents the fitted parameters for their associated attribute levels, reflecting participants’ preferences for referrals. The full model, incorporating all attributes, further details the interactions between the attributes or ASC and the personal and sociodemographic characteristics.

**Table 3.**
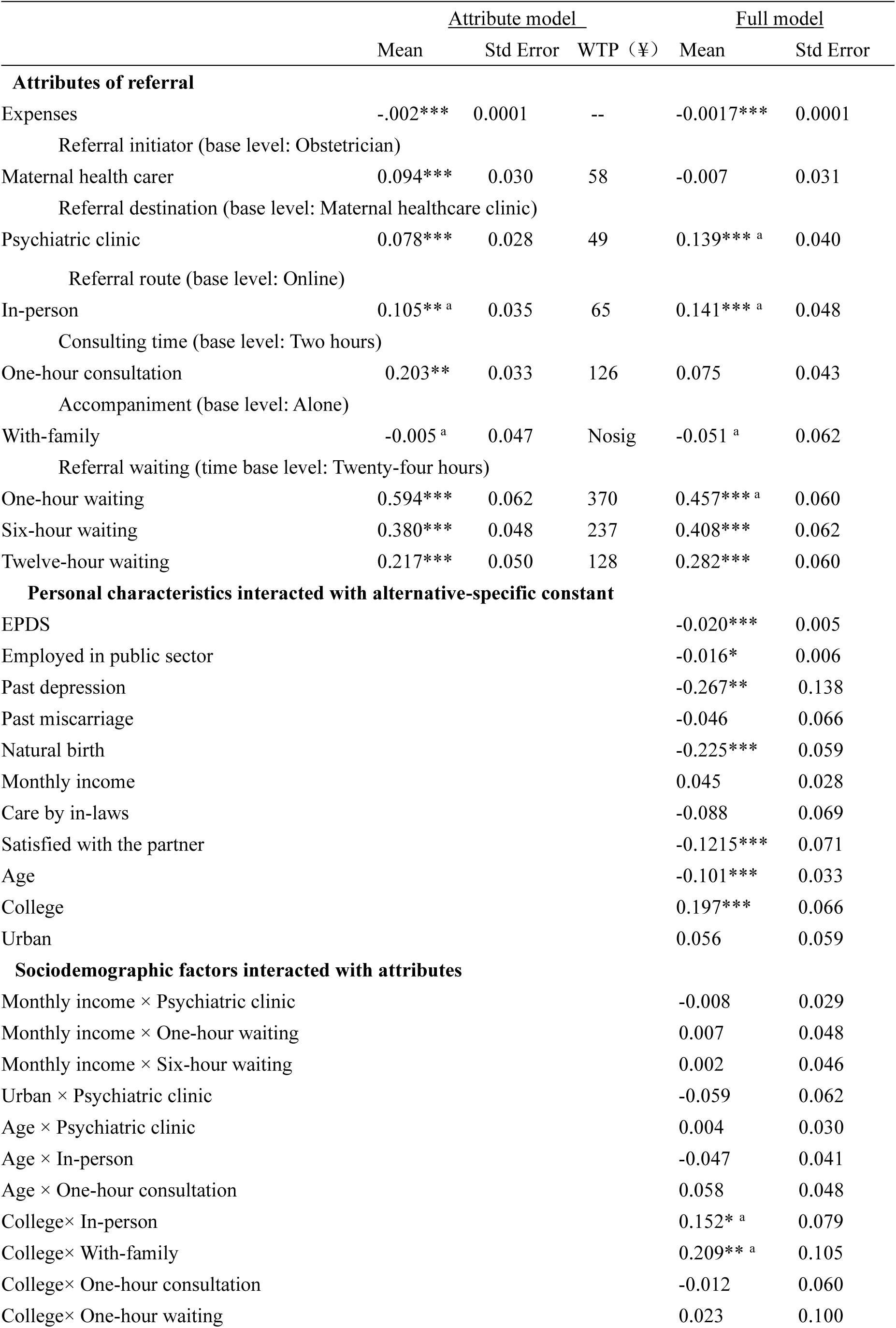
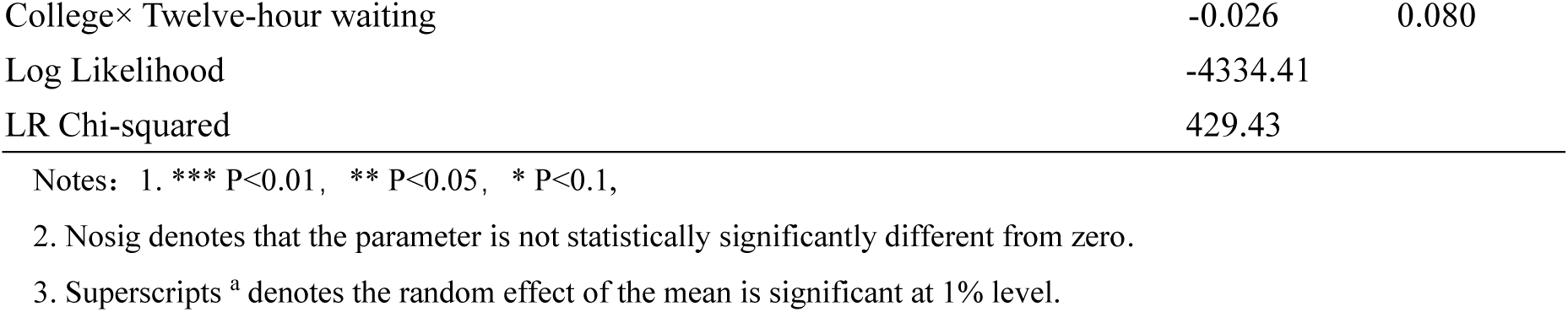
Results of mixed logit models and willingness to pay (WTP) for referral uptake.

|  | <u>Attribute model</u> |  |  | <u>Full model</u> |  |
| --- | --- | --- | --- | --- | --- |
|  | Mean | Std Error | WTP (¥) | Mean | Std Error |
| **Attributes of referral** |  |  |  |  |  |
| Expenses | -.002*** | 0.0001 | -- | -0.0017*** | 0.0001 |
| Referral initiator (base level: Obstetrician) |  |  |  |  |  |
| Maternal health carer | 0.094*** | 0.030 | 58 | -0.007 | 0.031 |
| Referral destination (base level: Maternal healthcare clinic) |  |  |  |  |  |
| Psychiatric clinic | 0.078*** | 0.028 | 49 | 0.139*** a | 0.040 |
| Referral route (base level: Online) |  |  |  |  |  |
| In-person | 0.105** a | 0.035 | 65 | 0.141*** a | 0.048 |
| Consulting time (base level: Two hours) |  |  |  |  |  |
| One-hour consultation | 0.203** | 0.033 | 126 | 0.075 | 0.043 |
| Accompaniment (base level: Alone) |  |  |  |  |  |
| With-family | -0.005 a | 0.047 | Nosig | -0.051 a | 0.062 |
| Referral waiting (time base level: Twenty-four hours) |  |  |  |  |  |
| One-hour waiting | 0.594*** | 0.062 | 370 | 0.457*** a | 0.060 |
| Six-hour waiting | 0.380*** | 0.048 | 237 | 0.408*** | 0.062 |
| Twelve-hour waiting | 0.217*** | 0.050 | 128 | 0.282*** | 0.060 |
| **Personal characteristics interacted with alternative-specific constant** |  |  |  |  |  |
| EPDS |  |  |  | -0.020*** | 0.005 |
| Employed in public sector |  |  |  | -0.016* | 0.006 |
| Past depression |  |  |  | -0.267** | 0.138 |
| Past miscarriage |  |  |  | -0.046 | 0.066 |
| Natural birth |  |  |  | -0.225*** | 0.059 |
| Monthly income |  |  |  | 0.045 | 0.028 |
| Care by in-laws |  |  |  | -0.088 | 0.069 |
| Satisfied with the partner |  |  |  | -0.1215*** | 0.071 |
| Age |  |  |  | -0.101*** | 0.033 |
| College |  |  |  | 0.197*** | 0.066 |
| Urban |  |  |  | 0.056 | 0.059 |
| **Sociodemographic factors interacted with attributes** |  |  |  |  |  |
| Monthly income × Psychiatric clinic |  |  |  | -0.008 | 0.029 |
| Monthly income × One-hour waiting |  |  |  | 0.007 | 0.048 |
| Monthly income × Six-hour waiting |  |  |  | 0.002 | 0.046 |
| Urban × Psychiatric clinic |  |  |  | -0.059 | 0.062 |
| Age × Psychiatric clinic |  |  |  | 0.004 | 0.030 |
| Age × In-person |  |  |  | -0.047 | 0.041 |
| Age × One-hour consultation |  |  |  | 0.058 | 0.048 |
| College × In-person |  |  |  | 0.152* a | 0.079 |
| College × With-family |  |  |  | 0.209** a | 0.105 |
| College × One-hour consultation |  |  |  | -0.012 | 0.060 |
| College × One-hour waiting |  |  |  | 0.023 | 0.100 |

|  |  |  |
| --- | --- | --- |
| College× Twelve-hour waiting | -0.026 | 0.080 |
| Log Likelihood | -4334.41 |  |
| LR Chi-squared | 429.43 |  |

Notes: 1. \*\*\* P<0.01, \*\* P<0.05, \* P<0.1,
2. Nosig denotes that the parameter is not statistically significantly different from zero.
3. Superscripts <sup>a</sup> denotes the random effect of the mean is significant at 1% level.

The Expenses attribute, representing the out-of-pocket costs associated with referral and treatment, was negative and highly significant (β=-0.002, P<0.001). This indicates that the probability of referral uptake decreases as costs increase. Moreover, time was identified as an important factor. Among the non-economic attributes, the shortest waiting time option (one hour) ranked as the most influential factor (β=0.594, P<0.001). This was followed by the shortest Consultation time of one hour (β=0.203, P<0.001), In-person referral route (β=0.105, P < 0.001), the maternal healthcare officer as the Referral initiator (β=0.094, P < 0.001) and psychiatric clinic as the Referral destination (β=0.078, P<0.001).

In the attribute model, generally, all PPD referral attributes were significant determinants, with the exception of the Maternal health officer and With-family attributes. When presented with various attributes, participants demonstrated a preference for referrals to psychiatric clinics, lower costs, shorter waiting times, and in-person treatment. The significant standard deviations of the random coefficients for these attributes indicated strong preference heterogeneity.

Examining the full model with interaction terms revealed preference heterogeneity through various interactions involving personal characteristics and socio-demographic variables. Higher EPDS scores and previous experience with depression decreased preference for referral. Similarly, having given birth naturally and being satisfied with one’s partner also reduced referral preference. Employment in the public sector and older age further reduced referral uptake. Conversely, possessing a bachelor’s degree or higher (College), which indicates a higher level of education, increased referral preference. Going through college also significantly increased the referral preference while associating with In-person and With-family treatment. Results also showed that these interactions exhibited significant preference heterogeneity, as evidenced by the significant standard deviations in the fitted random parameters.

### 3.3 Willingness to pay for attributes

Based on participants’ expressed preferences, we determined the expected monetary value associated with each referral attribute level, as shown in Table 3. For example, participants were willing to pay ¥128 more for a 12-hour referral interval, ¥237 more for a six-hour one, or ¥370 more for a one-hour referral, all in comparison to a 24-hour one. Similarly, they were willing to pay ¥126 more to reduce the length of consultation from two hours to one. There was also a preference for in-person treatment, with participants willing to allocate ¥65 more for it compared to online treatment. Nonetheless, the Referral initiator and Referral destination had a more marginal impact on decisions: participants were willing to pay only ¥58 more for referrals initiated by maternal healthcare providers over those by obstetricians, and a mere ¥49 more for a specialized referral to a psychiatric clinic as opposed to a maternal healthcare clinic.

### 3.4 Analysis of latent class model

The significance of random coefficients from the mixed logit models in Table 3 further suggests preference heterogeneity. For example, heterogeneity could come from different education levels as the interaction terms of In-person and With-family were significant. Therefore, latent class analysis (LCA) was implemented to further explore the preferences based upon the choice of attributes. A total of four latent classes were identified by using both the bootstrap likelihood ratio test (BLRT) and the Lo-Mendell-Rubin (LMR) test (p < 0.05). The full results based on the optimal number of four groups can be found in the Appendix. LCA models have shown that Class 2 was the largest in size representing 42% of the sample, and Class 4 was the smallest with 13.6% of sample. Classes 1 and 3 were similar in size with 23% and 21% of the sample, respectively.

To determine which attributes played the most important role in the referral preferences, we investigated the relative importance statistics (Determann et al., 2016; Maaya et al., 2018; Ride & Lancsar, 2016; Vermunt & Magidson, 2005). The importance of each attribute is its contribution to the utility function to the aggregate contribution from all the eight attributes. The relative importance statistics are calculated as the difference between parameters for levels within attributes multiplied by the attribute value of the largest and smallest level. Results on WTP for each attribute by class were reported in Table 4. Table 4 also includes the calculated relative importance statistics and the top three attributes ranked within the class. Moreover, Figures 4 and 5 illustrate the variation of relative importance of attributes across and within four classes, respectively.

**Figure 4.**
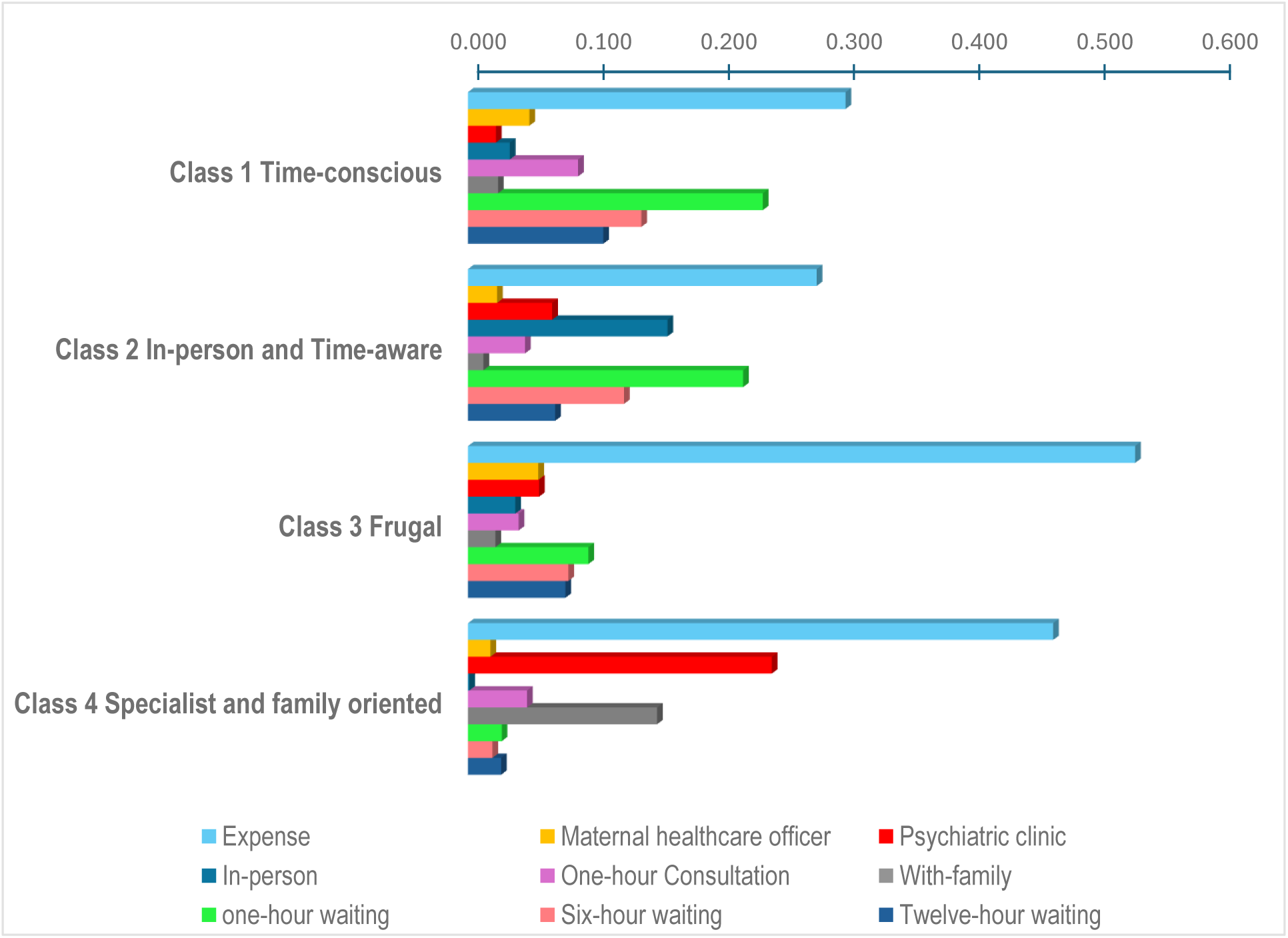
Relative importance of attributes by four classes

**Figure 5.**
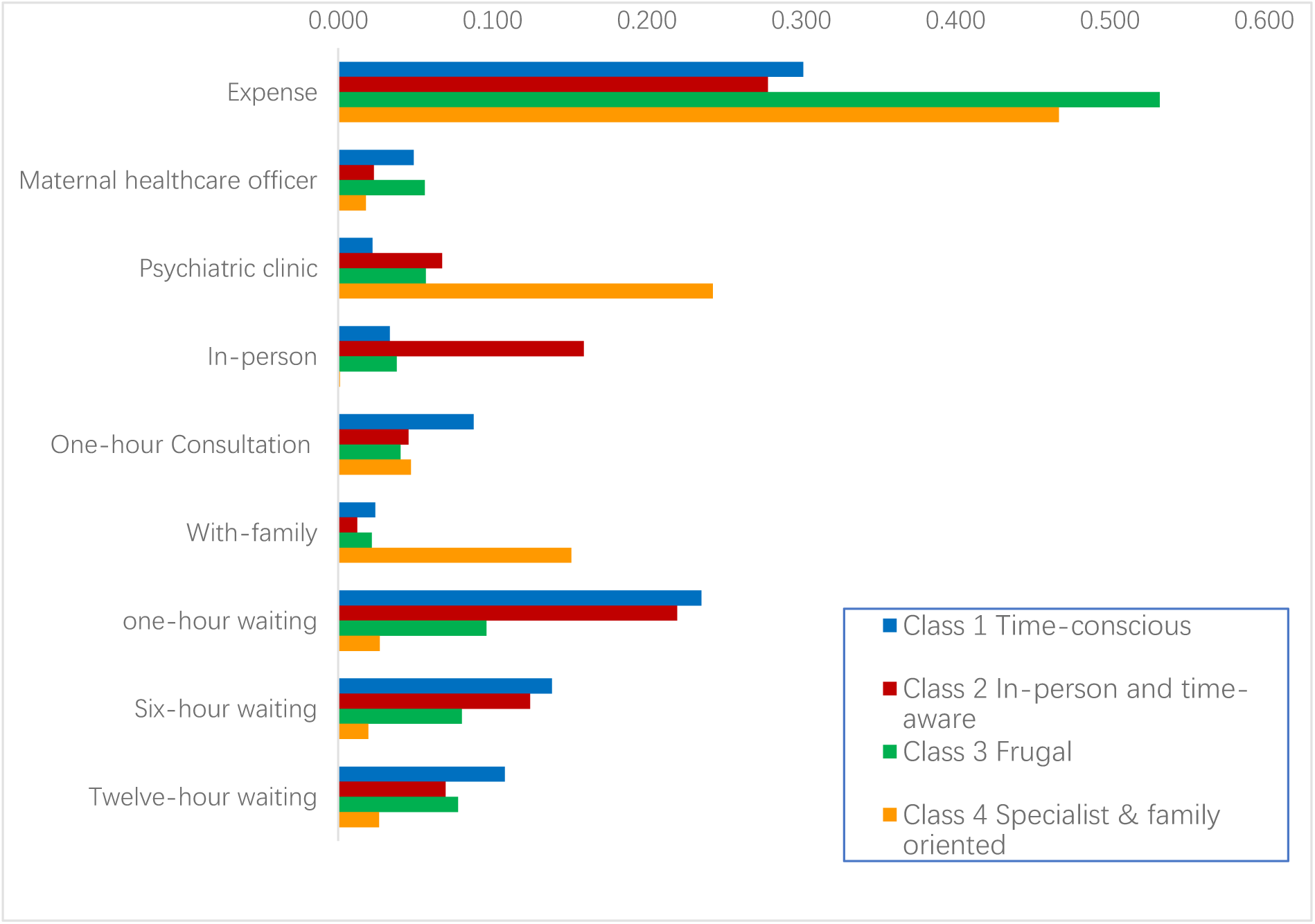
Relative importance by attributes

**Table 4.** Results of Relative importance statistics and WTP on attributes by latent classes.

|  | Class 1<br>Time-conscious | Class 2<br>Time-sensitive<br>and In-person | Class 3<br>Frugal | Class 4<br>Specialist &<br>family oriented |
| --- | --- | --- | --- | --- |
| Sample size (percentage) | 142 (23.1%) | 260 (42.1%) | 84 (13.6%) | 131 (21.2%) |
| Expense |  |  |  |  |
| [RIS] [rank] | **[0.303] [R1]** | **[0.278] [R1]** | **[0.532] [R1]** | **[0.467] [R1]** |
| Maternal healthcare officer |  |  |  |  |
| [RIS] [rank] | [0.049] | [0.023] | [0.056] | [0.018] |
| WTP | ¥110 | Nosig | ¥68 | Nosig |
| (LL,UL) | (72, 133) |  | (16, 122) |  |
| Psychiatric clinic |  |  |  |  |
| [RIS] | [0.022] | [0.067] | [0.057] | **[0.243] [R2]** |
| WTP | ¥49.6 | ¥136 | ¥69 | ¥349 |
| (LL,UL) | (8, 66) | (66., 209) | (16, 117) | (160, 527) |
| In-person |  |  |  |  |
| [RIS] (rank) | [0.144] | **[0.160] [R3]** | [0.038] | [0.001] |
| WTP | Nosig | ¥322 | Nosig | Nosig |
| (LL,UL) |  |  |  |  |
| One-hour consultation |  |  |  |  |
| [RIS] | [0.188] | [0.046] | [0.040] | [0.047] |
| WTP | ¥194 | Nosig | Nosig | Nosig |
| (LL,UL) | (145, 217) |  |  |  |
| With-family |  |  |  |  |
| [RIS] (rank) | [0.084] | [0.012] | [0.022] | **[0.151] [R3]** |
| WTP | [¥53] | Nosig | Nosig | [¥217] |
| (LL,UL) | (11, 70) |  |  | (45, 382) |
| One-hour waiting |  |  |  |  |
| [RIS] [rank] | **[0.235] [R2]** | **[0.219] [R2]** | [0.096] | [0.027] |
| WTP | ¥525 | ¥442 | Nosig | Nosig |
| (LL,UL) | (434, 570) | (270, 505) |  |  |
| Six-hour waiting |  |  |  |  |
| [RIS][rank] | **[0.138] [R3]** | [0.124] | [0.080] | [0.019] |
| WTP | ¥309 | ¥250 | Nosig | Nosig |
| (LL,UL) | (229, 339) | (176, 308) |  |  |
| Twelve-hour waiting |  |  |  |  |
| [RIS] | [0.108] | [0.070] | [0.078] | [0.026] |
| WTP | ¥242 | ¥150 | Nosig | Nosig |
| (LL,UL) | (190, 286) | (95, 172) |  |  |

Notes: 1. RIS denotes the relative importance statistics for each attribute. It is calculated as the difference of levels within parameters of attributes multiplied by the attribute value of the largest and smallest level. The maximum effect of each attribute is weighted across all the attributes. 2.. “Nosig” denotes that the parameter is not statistically significantly different from zero.

In general, the expense of the treatment is the top attribute for all the four groups as evidenced by the greatest relative importance statistics. Nevertheless, each of the four groups possessed their own characteristics while in favor of a lower-cost referral plan.

For Class 1, while most referral attributes evaluated in this study influenced this group’s decisions significantly, attributes pertaining to referral waiting time stood out. The relative importance statistics ranked the waiting time as the 2^nd^ highest referral attributes, except for expense. That is, this group expressed the strongest preference for plans that have a swift referral process. Their mean WTP for the shortest waiting time of one hour was as high as ¥525. We therefore labelled Class 1 as a “time-conscious” group.

Respondents in Class 2 were both time-sensitive and in-person oriented. This group considered attributes other than Referral waiting time and Accompaniment, supported by the top ranking in the relative importance statistics (RIS) of one-hour waiting time (RIS=0.219) and in-person (RIS=0.160). They favored in-person referrals to psychiatric clinics, and a shorter referral process. We labelled class 2 as “time-sensitive and in-person.”

Class 3 is much more cost-minded. This group primarily valued the attribute of expense, as the relative importance statistics reached 0.532, the highest among the four classes. They seemed to care about the Referral initiator (Maternal healthcare officer) and Referral destination (Psychiatric clinic), which were marginally significant. However, the relative importance statistics of Maternal healthcare officer and Psychiatric clinic and others were so low that we did not rank any other attributes for this group. We labelled this class as “frugal.”

The fourth class showed the most pronounced preferences for Referral destination (psychiatric clinic) and Accompaniment (with-family) according to the relative importance statistics. This pattern was also supported by the WTP estimates. This cohort of respondents assigned the highest WTP values among all classes for visiting a psychiatrist and wishing to go with family members. The WTP estimate for a referral to a psychiatric clinic stood out at ¥349, while the WTP to be accompanied by family members was ¥217. Thus, we labelled this class as “specialist-family oriented.”

Figure 4 illustrates the results of relative importance statistics in Table 4 for each attribute which varied from its base level for each of the four classes. The longest bar, ranking the highest out of all attributes for all classes, was “Expense”. The second most important attribute was the “one-hour waiting” time for Classes 1 and 2, visiting a “psychiatric clinic” for Class 4, while for Class 3, there was no clear attribute that stood out. There was no convergence regarding the ranking of the third most important attribute; each class varied.

From another perspective, each relevant importance statistics was summarized by attributes in Figure 5. Except for “Expense”, the last three clusters indicated that the referral waiting time (one-hour, six-hour, twelve-hour) emerged as the paramount non-economic attribute. This referral waiting time, preferred to be as short as an hour, comprises the entire process: starting from agreeing to a referral to reaching the referral destination, attending the appointment, and any waiting period involved.

More specifically, regarding whether and how the personal characteristics affected the latent classes, we decided to opt for the pairwise t-test. Results of mean comparisons were presented in Table 5. From Table 5, we observed some interesting findings among six pairwise t-tests. In general, nine out of eleven personal characteristics were statistically significant. Living in an urban area was found to be significantly different among three pairwise mean comparisons: Classes 1 vs 2, Classes 1 vs 4 and Classes 3 vs 4; this was followed by “College”, “Satisfied with partner” and “Past miscarriage” which were significant among two mean comparisons; then “EPDS”, “Employed in public sector”, “Monthly income”, and “Care by In-laws” were statistically different when making one pairwise mean comparison. The results also supported that the differences between latent classes could be explained by their associated personal characteristics. For example, the variation in preferences between Class 1 and 4 could also be driven by the significant differences in “Urban”, “Satisfied with partner”, “Monthly income”, “Past miscarriage” and “EPDS” in addition to effects of the attributes. On the other hand, the difference between Classes 1 and 3 appear to be solely driven by the referral attributes as there was no significance found in all six pair-wise mean comparisons.

**Table 5.** Results of mean comparison of personal characteristics by four latent classes.

|  | Class 1<br>Mean | Class 2<br>Mean | Class 3<br>Mean | Class 4<br>Mean | Class 1 vs<br>Class 2 | Class 1 vs<br>Class 3 | Class 1 vs<br>Class 4 | Class 2 vs<br>Class 3 | Class 2 vs<br>Class 4 | Class 3 vs<br>Class 4 |
| --- | --- | --- | --- | --- | --- | --- | --- | --- | --- | --- |
| **EPDS** | 12.64 | 13.12 | 13.88 | 14.08 | 0.471 | 0.168 | **0.053\*** | 0.371 | 0.162 | 0.830 |
| **Employed in public sector** | 0.37 | 0.31 | 0.44 | 0.36 | 0.265 | 0.238 | 0.911 | **0.030\*** | 0.302 | 0.194 |
| Past depression | 0.03 | 0.03 | 0.05 | 0.06 | 0.885 | 0.563 | 0.180 | 0.614 | 0.175 | 0.604 |
| **Past miscarriage** | 0.33 | 0.25 | 0.31 | 0.25 | **0.060\*** | 0.798 | **0.078\*** | 0.215 | 0.887 | 0.284 |
| Natural birth | 0.62 | 0.54 | 0.59 | 0.56 | 0.117 | 0.671 | 0.234 | 0.420 | 0.769 | 0.582 |
| **Monthly income** | 6701 | 7018 | 6860 | 7453 | 0.396 | 0.751 | **0.063\*** | 0.733 | 0.227 | 0.225 |
| **Care by In-laws** | 0.21 | 0.23 | 0.15 | 0.25 | 0.603 | 0.329 | 0.359 | 0.142 | 0.623 | **0.079\*** |
| **Satisfied with the partner** | 0.78 | 0.87 | 0.85 | 0.90 | **0.016\*\*** | 0.160 | **0.003\*\*** | 0.663 | 0.405 | 0.305 |
| **Age** | 30.47 | 30.03 | 30.49 | 30.85 | 0.314 | 0.978 | 0.427 | 0.401 | **0.053\*** | 0.530 |
| **Age ( > 30)** | 0.52 | 0.51 | 0.55 | 0.58 | 0.747 | 0.710 | 0.243 | 0.510 | **0.100\*** | 0.559 |
| **College** | 0.32 | 0.47 | 0.41 | 0.35 | **0.001\*\*\*** | 0.165 | 0.531 | 0.284 | **0.009\*\*** | 0.368 |
| **Urban** | 0.65 | 0.74 | 0.67 | 0.81 | **0.031\*\*** | 0.705 | **0.001\*\*\*** | 0.183 | 0.146 | **0.020\*\*** |
| Sample size | 142 | 260 | 84 | 131 |  |  |  |  |  |  |

. Notes: Mean comparison of personal characteristics across the four classes. Pairwise p-value from the t-tests on the equality of means among classes 1–4. The significance of the tests is indicated with \*p < 0.1, \*\*p < 0.05 or \*\*\*p < 0.01.

## 4. Discussion

Our study investigated women’s preferences for the uptake of referral for PPD. It analyzed the various attributes of referral and women’s personal and demographic characteristics and found evidence for the existence of significant heterogeneity. Our study also provided quantitative analysis of the importance of the attributes after latent class analysis.

The expense was the most decisive characteristic for women to decide whether to opt for a referral or not. In addition to this, all time-related attributes were also key factors for taking a referral. Waiting times of three levels (one-, six- and twelve-hour), and even the consultation time, all ranked relatively highly and had high WTP values. The respondents favored in-person treatment as the referral route, followed by referral initiated by their maternal health officer, then choosing psychiatric clinics as the referral destination. The only attribute that had no significant effect was the attribute of Accompaniment; in general, respondents showed no preference for visiting the consultation clinic by themselves or with family members. Respondents decreased their preferences in referral uptake under various personal circumstances: having a higher EPDS score, working in the public sector, having previously experienced depression, giving birth naturally, being older, and being satisfied with their partners. On the other hand, having a bachelor’s degree or higher was found to increase their preference for referral uptake.

Notably, distinct heterogeneities in preferences were observed from the results of the mixed logit models — not only through the attributes, but also through a set of personal characteristics. Subsequent latent class analysis revealed that participants could be categorized into four classes, namely “time-conscious” (Class 1), “time-sensitive and in-person” (Class 2), “frugal” (Class 3), and “specialist and family-oriented” (Class 4), based on their unique characteristics. Personal characteristics and socio-demographic information differed significantly across the four latent classes. Because Class 2 alone represented 42% of the respondents and Classes 1, 2 and 4 together made up 86% of the sample, it was highly likely that we found a general preference model for PPD referral.

### 4.1 Understanding the DCE results: insights for policy and practice

In terms of economic attributes, all participants preferred low-cost referral plans. This aligns with findings from previous studies that highlight cost as a paramount factor influencing patients’ healthcare decisions (J. J. Kim et al., 2010; Watkins et al., 2014). Particularly, the result was consistent with Ride and Lancsar (2016), who found that the cost barrier is a significant factor influencing treatment plans for perinatal depression and anxiety (Ride & Lancsar, 2016). However, it is worth noting that researchers generally discourage drastic cost reductions (Si et al., 2019; Sun et al., 2019; Wong et al., 2020; Zhou et al., 2015; Zhu et al., 2019). Such caution comes from considerations regarding the marginal effect and practical challenges in implementation, such as the potential inability to offer adequate physician compensation. Given this, it becomes vital to pinpoint and employ non-economic strategies that also significantly drive referral uptake.

Possible explanations for the dominant importance of the time attribute could be concern on the part of the respondents who were all mothers of newborns, given potential inconvenience and inaccessibility of the referral system. Drawing a broad reference, free childcare was found as an incentive to choose a perinatal depression and anxiety (PNDA) treatment, while breastfeeding discouraged new mothers from taking up PNDA treatment (Ride & Lancsar, 2016).

All four participant groups showed a marked preference for a referral destination of psychiatric clinics. Importantly, these groups did not shy away from consulting a doctor due to stigma or embarrassment, contrary to insights from earlier qualitative studies (Byatt et al., 2013; Flynn et al., 2010) and the most recent reports by Xue (2023) and (W. Xue et al., 2023). On the downside, the allocation and distribution of mental health resources in China leave much to be desired. Most accessible psychiatric clinics are clustered within regional medical centers. This centralized distribution creates large disparities in mental health services, posing significant challenges in meeting the needs of new mothers (Chen et al., 2020; Shi et al., 2019).

Interestingly, participants preferred maternal healthcare providers, rather than obstetricians, to initiate referrals. Existing qualitative research typically presumes obstetricians play this pivotal role (Bayrampour et al., 2018; Byatt et al., 2012; Elkhodr et al., 2018; Fletcher et al., 2019; Gärtner et al., 2015; H. Y. Kim et al., 2019; Larson et al., 2015). However, within China’s distinct maternal and child healthcare framework, maternal healthcare providers interact more frequently with pregnant women, from the first trimester through six weeks post-delivery. The rapport between these healthcare providers and their patients, along with the professional’s demeanor, heavily influence a pregnant woman’s decision to accept a referral (Byatt et al., 2013; Canty et al., 2019; Dennis & Chung-Lee, 2006; Elkhodr et al., 2018; Forde et al., 2020; W. Xue et al., 2023). Accordingly, when establishing a PPD referral mechanism, it is essential to prioritize service accessibility and convenience, while also addressing economic concerns.

The results further indicated the existence of preference heterogeneity in the referral route and revealed four distinct participant types. The “time-conscious” (Class 1) preference for online referrals aligns with the finding of a higher acceptance rate of online diagnosis and treatment schemes among PPD patients (Gong et al., 2020). However, it differed from the finding by Ride and Lancsar (Ride & Lancsar, 2016), of no effect of accepting an online treatment. In contrast, the largest class of “time-sensitive and in-person” (Class 2) which represented 42.1% of respondents, was more inclined towards direct, face-to-face medical consultations, possibly because psychological therapy is often more well-received when conducted in person (Ride & Lancsar, 2016).

Family played an important role as well. As indicated by the Accompaniment attribute, both the “time-conscious” (Class 1) and “specialist and family-oriented” (Class 2) groups, constituting 44.3% of participants, expressed a desire for family accompaniment during their appointment. In the mixed logit model, the participants who were satisfied with their partners were less likely to take the referral. Our findings provided supporting evidence for other studies (Cox et al., 2017; Goodman, 2009) as depression can often deter individuals from seeking healthcare services (Maneze et al., 2016). Consequently, social support serves as a pivotal moderator. While improving social support can stimulate patients with depression to seek assistance proactively, a gap in understanding on the family’s part can act as a significant barrier (Noonan et al., 2017; Sword et al., 2008). Hence, merely focusing on maternal mental health is not sufficient. A more holistic approach necessitates extending mental health education to include the patient’s immediate social circle, such as relatives and friends.

### 4.2 Comparison with findings in the literature

While other choice studies explored preferences for treatment of perinatal depression and anxiety (Ride and Lancsar, 2016), or anxiety and depression in general (Muntingh et al. 2019), to our knowledge, our study is the first DCE to examine the relationship between the referral process and women’s preferences. In our study, we found that respondents decreased their preferences for referral when they were happy with their partner, which broadly supports the findings by Abe-Kim et al (2002) and Kakuma et al (2011)(Abe-Kim et al., 2002; Kakuma et al., 2011). These two studies found evidence that family support is a key factor in not using mental health and medical services.

In addition to finding combinations of preferred attributes, our study further quantified the relative importance of the attributes by comparing the relative importance statistics and WTP estimates. We grouped the respondents into four classes and compared the characteristics of each, and although we could not directly compare our results to other PPD referral studies, we have drawn broad references to related choice studies, especially on depression and anxiety.

## 5. Limitations

There are several limitations of our study. Firstly, while a meta-analysis has shown that DCEs can provide a reasonable prediction of health-related behaviors,^63^ in our current study we did not consider an option of “no referral needed” in our DCE scenarios. This was because we wanted to focus on analysing the preferences for choosing referrals. It would also be interesting to explore the specific reasons why mothers with PPD did not seek treatment in the future. Doing so would provide us with more information on how to implement early intervention.

Secondly, due to local COVID-19 policies, we were unable to conduct the planned qualitative research in Changsha to obtain DCE attributes. We instead sourced these attributes from analogous regions, using guidance from expert consultations and qualitative interviews.

Thirdly, even though we employed stratified cluster random sampling, our final sample had slightly disproportionate representation of urban residents.

Lastly, our target population for the study consisted of new mothers who screened positive for PPD. This specific focus made it challenging to amass a large sample. While we met the minimal sample size required for statistical analysis, the individual group sizes in the post-latent class analysis were only moderate.

## 6. Conclusion

To the best of our knowledge, this is the first stated-preference study using discrete choice experiment to examine the referral uptake of postpartum depression. In general, the factors of time, referral initiator, referral destination and cost were found to be pivotal factors in referral uptake. Furthermore, this study found significant heterogeneity in the new mothers’ referral preferences while using mixed logit models, and further categorized participants into four groups. To improve uptake of referral for postpartum depression, it would be beneficial to introduce more flexibility in cost-effective referral programs to meet the varied preferences among patients.

## Data Availability

All data produced in the present study are available upon reasonable request to the authors.

## Appendix A Table Results of t-test comparisons

| Variable | Mean (std dev) | t statistic (p value) |
| --- | --- | --- |
| Age | 30.32 (4.39) | 1.652 (0.11) |
| Completion of college:<br>yes | 0.41 (0.49) | -0.513 (0.608) |
| Employed: yes | 0.45 (0.50) | 0.618 (0.537) |
| Monthly income (¥) | 7081 (3756) | -0.805 (0.421) |
| Income decrease: yes | 0.56 (0.50) | -1.940 (0.053) |
| Complications: yes | 0.20 (0.40) | -0.994 (0.321) |
| Delivery method: natural<br>birth | 0.571 (0.495) | 0.191 (0.849) |
| Miscarriage: yes | 0.267 (0.443) | 1.208 (0.228) |

## Appendix B Table Results of goodness of fit from latent class analysis

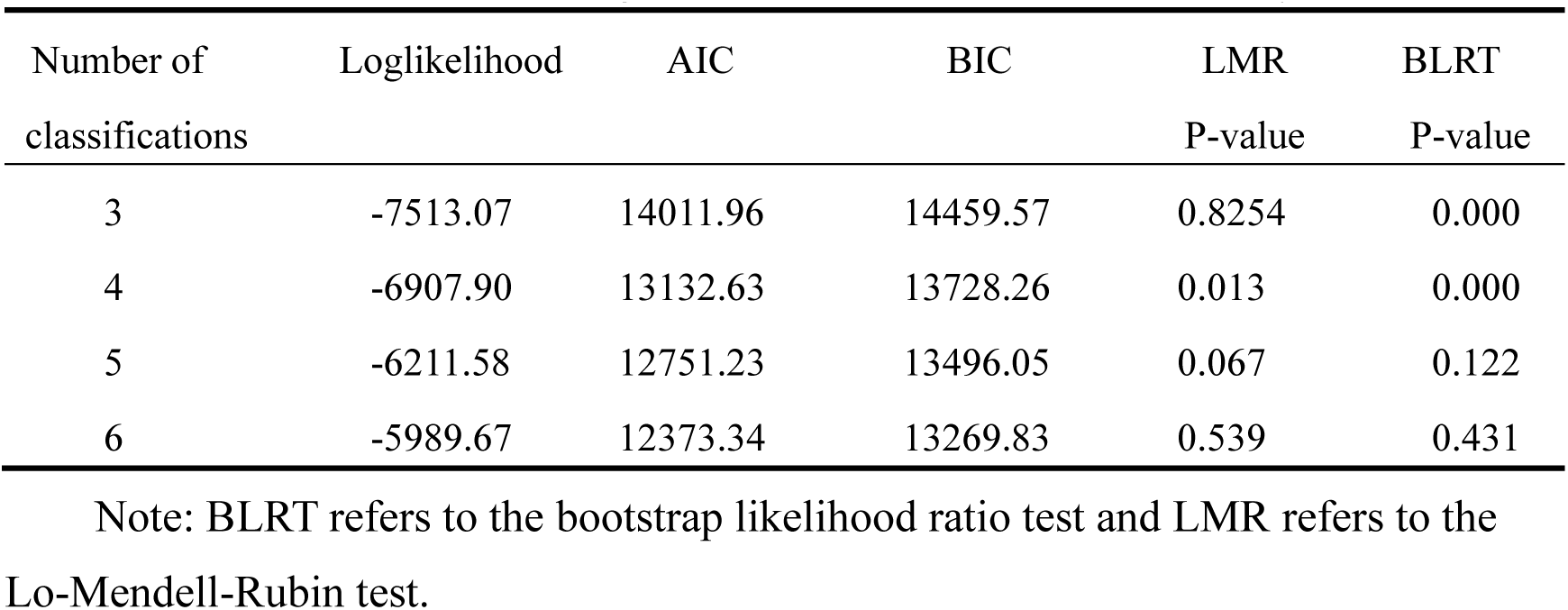

| Number of<br>classifications | Loglikelihood | AIC | BIC | LMR<br>P-value | BLRT<br>P-value |
| --- | --- | --- | --- | --- | --- |
| 3 | -7513.07 | 14011.96 | 14459.57 | 0.8254 | 0.000 |
| 4 | -6907.90 | 13132.63 | 13728.26 | 0.013 | 0.000 |
| 5 | -6211.58 | 12751.23 | 13496.05 | 0.067 | 0.122 |
| 6 | -5989.67 | 12373.34 | 13269.83 | 0.539 | 0.431 |

Note: BLRT refers to the bootstrap likelihood ratio test and LMR refers to the Lo-Mendell-Rubin test.

## Appendix C Results from unconstraint logit model for latent classes

|  | Time-sensitive | In-person | Frugal | Specialist-oriented |
| --- | --- | --- | --- | --- |
| Sample size<br>(percentage) | 142<br>(23.1%) | 260 (42.1%) | 84 (13.6%) | 131 (21.2%) |
| Expense | -0.003*** | -0.0006*** | -0.003*** | -0.003*** |
| Referral initiator:<br>Maternal healthcare officer | 0.293*** | -0.030 | 0.190* | -0.069 |
| Referral destination:<br>Psychiatric clinic | 0.133* | 0.087** | 0.192* | 0.935*** |
| Referral route: Offline | -0.200*** | 0.206** | 0.128 | -0.004 |
| Consultation length:<br>One hour | 0.525*** | 0.059 | 0.137 | 0.182 |
| Accompaniment:<br>Accompanied by family members | 0.143** | -0.016 | -0.074 | 0.582*** |
| Interval time:<br>One hour | 1.406*** | 0.284*** | 0.325 | 0.104 |
| Six hours | 0.827*** | 0.161*** | 0.271 | 0.075 |
| Twelve hours | 0.645*** | 0.090** | 0.263 | 0.102 |

Note: \*\*\* P<0.01, \*\* P<0.05, \* P<0.10

## Notes

### Competing Interest Statement

The authors have declared no competing interest.

### Funding Statement

This study did not receive any funding.

### Author Declarations

IRB of Central South University gave ethical approval for this work(CTXY-2020-102).

